# Protocol for a prospective, hospital-based registry of pregnant women with SARS-CoV-2 infection in India: PregCovid Registry study

**DOI:** 10.1101/2021.07.21.21260823

**Authors:** Rahul K Gajbhiye, Niraj N Mahajan, Rakesh Waghmare, Suchitra Surve, Prashant Howal, Aishwarya Bhurke, Merlin Pious, Deepak N Modi, Smita D Mahale

**Affiliations:** Clinical Research Lab, ICMR-National Institute for Research in Reproductive Health, Mumbai, India; Department of Obstetrics & Gynecology, Topiwala National Medical College & BYL Nair Charitable Hospital, Mumbai India; Medical Education and Drugs Department, Government of Maharashtra, India; Department of Community Medicine, Grant Government Medical College, and Sir J. J. Group of Hospitals, Mumbai, India; Molecular and Cellular Biology Laboratory, ICMR-National Institute for Research in Reproductive Health, J M Street, Parel, Mumbai; Structural Biology Department, ICMR-National Institute for Research in Reproductive Health, J M Street, Parel, Mumbai, India

**Keywords:** COVID-19, Coronaviruses, New-born, Pregnancy, Registry, Viruses, Vertical transmission

## Abstract

**Introduction:** Pregnant women are at increased risk of contracting coronavirus disease 2019 (COVID-19) due to several factors and therefore require special attention. However, the consequences of the COVID-19 pandemic on pregnant women and their newborns remain uncharted. The PregCovid registry aims to document the impact of SARS-CoV-2 infection on pregnant, post-partum women and their newborns. The aim of the registry is also to determine mother-to-child transmission of SARS-CoV-2 infection in India.

**Methods and analysis:** PregCovid is a hospital-based registry for capturing information of pregnant, post-partum women with COVID-19 and their newborns in India. Medical case records of pregnant and post-partum women with laboratory-confirmed diagnoses of COVID-19 will be captured in real-time using an online electronic patient record (EPR) software. The frequency of each symptom will be calculated. The laboratory data will be analyzed for calculating the frequency of laboratory parameters consistently higher in women with COVID- The adverse pregnancy and neonatal outcomes will be analyzed and their frequency will be calculated. Response to treatment will be analyzed for frequency calculation (number of women treated with different treatment regimens). The mother-to-child transmission data will be analyzed from the RT-PCR and/ antibody data of neonatal and maternal samples tested wherever the information is available. The registry data will be crucial for developing strategies for reducing the adverse impact of COVID-19 on pregnant women and their new-born.

**Ethics and dissemination:** The study is approved by the Institutional Ethics Committee of ICMR-National Institute for Research in Reproductive Health (#55/2020), BYL Nair Hospital, Mumbai, India (# 63/2020); and all the 18 participating study sites under Medical Education and Drugs Department of Government of Maharashtra. The Institutional Ethics Committees granted a waiver of consent as the data is collected from the medical case records.

**Trial registration number:** CTRI/2020/05/025423

**Article summary:** *Strengths and limitations of this study:*

- The PregCovid registry is a hospital-based registry at dedicated COVID-19 hospitals in India. The registry will help to identify new epidemiological, clinical characteristics, obstetrics outcomes associated with pregnant women and/post-partum women with COVID-19 in India. The study will also generate information on clinical presentations and outcomes of neonatal born to mothers with COVID-19 in India.
- PregCovid registry will provide evidence of mother to child transmission of SARS-CoV-2 infection in Indian women
- The evidence on unusual presentations of COVID-19 in pregnant and post-partum women will be generated.
- The follow-up of participants is only till the discharge from the hospital. Long-term follow-up is not included in the study.

## Introduction

Coronavirus disease 2019 (COVID-19) has emerged as a public health emergency with more than 103 million confirmed cases of COVID-19, including 2.3 million deaths, worldwide as [1]. Several variants of SARS-CoV-2 are now emerging [2] however, it is still unknown whether they have any clinical impact on pregnant women. Several International and National agencies are actively engaged to address the impact of the COVID-19 pandemic. Previously, members of the coronavirus family such as severe acute respiratory syndrome coronavirus (SARS-Co-V) and Middle East Respiratory Syndrome (MERS-CoV) were reported to be associated with severe complications during pregnancy like miscarriage, fetal growth restriction, preterm birth, and maternal deaths [3]. In general, pregnant women are a vulnerable population and need special attention. Pregnant women are particularly susceptible to respiratory pathogens and severe pneumonia, due to various factors such as physiologic changes in the immune and cardiopulmonary systems (e.g. diaphragm elevation, increased oxygen consumption, and edema of respiratory tract mucosa) which make them at risk of hypoxia [4]. Since the initial period of reporting case reports or case series on pregnant women with COVID-19 from China, now there are several studies published from different geographical regions. Initial studies mainly from China reported that clinical presentation of pregnant women with COVID-19 was comparable to non-pregnant cases and there was no risk of mother to child transmission of Severe acute respiratory syndrome coronavirus 2 (SARS-CoV-2) infection [5]. However, subsequently, studies were demonstrating the adverse impact of COVID-19 on both maternal and newborn health [4,6–9]. Evidence is gathering which suggests that there might be population biases in the susceptibility of COVID-19. Analysis of 427 pregnant women of the UK registry revealed that there might be racial differences in the severity of presentation and outcomes [7]. Also, a higher incidence of maternal death due to COVID-19 is reported in some countries as compared to China or Europe [10]. Similarly, in the United States of America (USA), ethnic disparities in incidence and outcomes are also observed in non-pregnant populations with COVID-19, notably in the USA [11]. Factors such as social behavior, health-seeking behavior, comorbidities, and unknown genetic influences could be possible causes of differences in presentation and outcomes of COVID-19. This highlights a need for population-specific data of pregnant women, to gain an insight into social-epidemiological and clinical determinants of outcomes in COVID-19.

Currently, in India, the number of COVID-19 cases is on the rise. The Ministry of Health and Family Welfare (MoHFW), Government of India, reported 1,04,80,455 total confirmed cases with 1,54,703 deaths as of 4^th^ February 2021 [12]. Out of these, Maharashtra state contributed 19,43,335 cases with 51,169 deaths due to COVID-19 [12]. COVID-19 affected Indian states including Maharashtra which need special attention in planning the strategies for combating COVID-19, especially in the vulnerable population such as pregnant women. Our preliminary observations suggest that nearly 12% of pregnant women in Maharashtra State have SARS-CoV-2 infection and about 10% of these are symptomatic, highlighting the emergency nature of the situation [13]. Currently, there is no epidemiological, demographic, and clinical information on pregnant women with COVID-19 in India. Additionally, health outcomes of neonates born to COVID-19 mothers have not been documented in the Indian population.

To address the knowledge gaps, ICMR-National Institute for Research in Reproductive Health (NIRRH) has initiated a hospital-based pregnancy registry to capture socio-demographic, clinical presentations, treatment outcomes, obstetric and neonatal outcomes in Indian women with COVID-19. Herein, we describe the protocol and the characteristics of this registry.

### Aim of PregCovid study

The following research questions will be addressed:

1. What are the clinical characteristics of pregnant and post-partum women who are diagnosed with SARS-CoV-2 infection?
2. What are the pregnancy outcomes in Indian women with SARS-CoV-2 infection?
3. How does the treatment of SARS-CoV-2 infection in pregnancy influence the outcomes for mother and infant?
4. Are there any ethnic or socioeconomic determinants of clinical presentations and outcomes of COVID-19 in pregnant women?

### Objectives

The objectives of the study are: a) To understand the socio-demographic, clinical presentations, reproductive characteristics of pregnant and post-partum women diagnosed with SARS-CoV-2 infection b) To determine the pregnancy and fetal outcomes in women with SARS-CoV-2 infection and study the effect of treatment on pregnancy outcomes c) To evaluate the proportion of maternal to the fetal transmission of SARS-CoV-2 infection in pregnancy.

## Materials and methods

The PregCovid registry is a prospective, hospital-based study designed to capture hospital data of pregnant and post-partum women with COVID-19 and their newborns.

### Identification of study sites for data capturing

Since Maharashtra is a hotspot region for COVID-19, it was decided initially to launch the registry to capture the data in the state. Towards that, the Medical Education and Drugs Department (MEDD) Government of Maharashtra and Municipal Corporation of Greater Mumbai (MCGM) were consulted, and primary data on COVID-19 and pregnant women were collected by a rapid survey.

### Data collection instrument

An *a priori* requirement of the registry was the development of the case record form. Towards this RG and DM carried out a situational analysis of the current evidence on the effectiveness and outcomes of COVID-19 on pregnant women [14]. Based on the outcomes of the systematic review of the baseline data of more than 400 women from different parts of the world and also reviewing the USA registry PRIORITY: Pregnancy Coronavirus Outcomes Registry [15], we shortlisted the key parameters about which the information should be collected.

In the next step, the case record form was independently reviewed by a team of gynecologists and pediatricians. Their inputs were recorded and the case record form was modified by a consensus. The modified case record form was circulated to the Investigators (Professor & Head, Obstetrics and Gynecology Departments, and Professor & Head Pediatrics/Neonatology Department) of the PregCovid Registry Network for further feedback and its feasibility as they have experience of treating the COVID-19 cases. RG coordinated with Investigators of the PregCovid Registry Network Hospitals for pilot testing of the case record form. Based on the feasibility and pilot testing, the case record form was modified and finalized by the consensus of investigators (Table 1).

**Table 1:** Maternal and New-born information concerning different parameters captured by Electronic Patient Record for PregCovid registry

| Maternal Information | Parameters |
| --- | --- |
| **Socio-Demographic and Epidemiological data** | Age, Religion, Occupation, Education, Financial status,<br>Residential area (Red zone, Orange, Green) |
| **Medical History** | Anemia, Diabetes Mellitus, Chronic Hypertension, Blood disorders, Chronic obstructive disease, Asthma, Tuberculosis, Cardiac disorders, Psychiatric disorders, Thyroid disorders, Kidney disorder, rheumatoid arthritis, malignancy, Immunocompromised state, Previous surgical history, H/o contact in the community, exposure while caring for COVID-19 patient and foreign travel |
| **Obstetric History** | Gestational Age, and conception history, Gravida, Parity, Mode of delivery, Living Children, Abortions, Stillbirth, Ectopic Pregnancy, Molar Pregnancy, Preterm Birth |
| **Clinical Presentation of COVID-19** | Asymptomatic or Symptomatic, details of symptoms, Duration of symptoms, Intervals–symptoms onset to admission, Exposure/contact to diagnosis, Atypical Symptoms, Complications of COVID-19 (ARDS, Consolidation, Heart Failure, Septic Shock, Acute Renal Injury, etc.) |
| **General Physical Examination** | BMI, Stability of vitals, Oxygen Saturation at time of Admission, Lowest Oxygen saturation during hospitalization, Date, and method of COVID-19 Testing, Type of sample for COVID-19 Testing, Indications of COVID-19 Testing, X-ray, CT scan, other Laboratory Investigations, Co-infections like Malaria, Dengue, etc |
| **Delivery Details** | *Pregnancy Outcomes:* Delivered (Singleton/Multifetal), Undelivered, Spontaneous Abortion, Stillbirth, Ectopic Pregnancy, MTP<br>*Delivery details:* Mode of delivery, Duration of labor, Induction, CS details, Complications<br>*Pregnancy Complications:* Preeclampsia, Eclampsia, Placenta Previa, Placenta Abruption, Uterine Rupture, Premature Rupture Of Membranes, Oligohydramnios, Polyhydramnios, Antepartum |

|  | Postpartum Hemorrhage, Gestational Hypertension, Gestational Diabetes/ Pregestational Diabetes Mellitus, Preterm Labour, Chorioamnionitis, Fetal Growth Restriction, Congenital Malformation, Hematoma, Surgical Site Infection |
| --- | --- |
| **Treatment** | Antibiotics, Antivirals, Hydroxychloroquine, Steroids, Heparin, High-flow nasal cannula oxygen therapy, NIV, any other treatment and outcomes, Hospital and ICU stay, Blood transfusion, Convalescent Plasma Therapy |
| Newborn Information | Parameters |
| **Basic demographic and clinical investigations** | Date of birth, 5 min APGAR score, Gender, Congenital anomalies, Birth Weight, Feeding Details, NICU admission, Neonatal Complications, Indications and duration of NICU admission, Vaccination details, Testing for COVID-19, Cord blood testing for COVID-19( if available), Laboratory Investigations, Treatment and outcomes. |

*APGAR- Appearance, Pulse, Grimace, Activity, and Respiration, CS- Cesarean delivery, MTP- Medical Termination of Pregnancy, NIV- Non-Invasive Ventilation*

### Tools for real-time data capture

To capture the real-time data, an online electronic patient record (EPR) software was developed by an outsourced team of experts. The developed software was first tested using dummy data by a team of obstetricians and pediatricians from the participating study sites. The time for data entry was recorded for obstetrics and pediatrics sections. Based on the inputs received from the user team, the software was modified and pilot testing was carried out before the actual start of the data entered into the software.

### Approval from Institutional Review Boards (IRB)

The study is approved by the Institutional Ethics Committee of ICMR-National Institute for Research in Reproductive Health (#55/2020), BYL Nair Hospital, Mumbai, India (# 63/2020); and all the 18 participating study sites under Medical Education and Drugs Department of Government of Maharashtra. The Institutional Ethics Committees granted a waiver of consent as the data is collected from the medical case records.

### CTRI Registration

The study is registered with the Clinical Trials Registry, India (CTRI) and the registration number is CTRI/2020/05/025423.

### Characteristics of the PregCovid registry network and requirements of additional participating centers

The overall flow of the study protocol is outlined in Figure 1. The registry is presently initiated with 19 participating centers; it is open for all public and private COVID-19 hospitals all over India. A dedicated PregCovid portal for data entry (http://app.pregcovid.com/login) is developed along with a website for giving information about the registry for prospective collaborators, publications, and any breakthrough information related to the registry (https://pregcovid.com/). BYL Nair Hospital, Mumbai, a dedicated COVID-19 hospital was the first study site of PregCovid registry [16].

**Figure 1:**
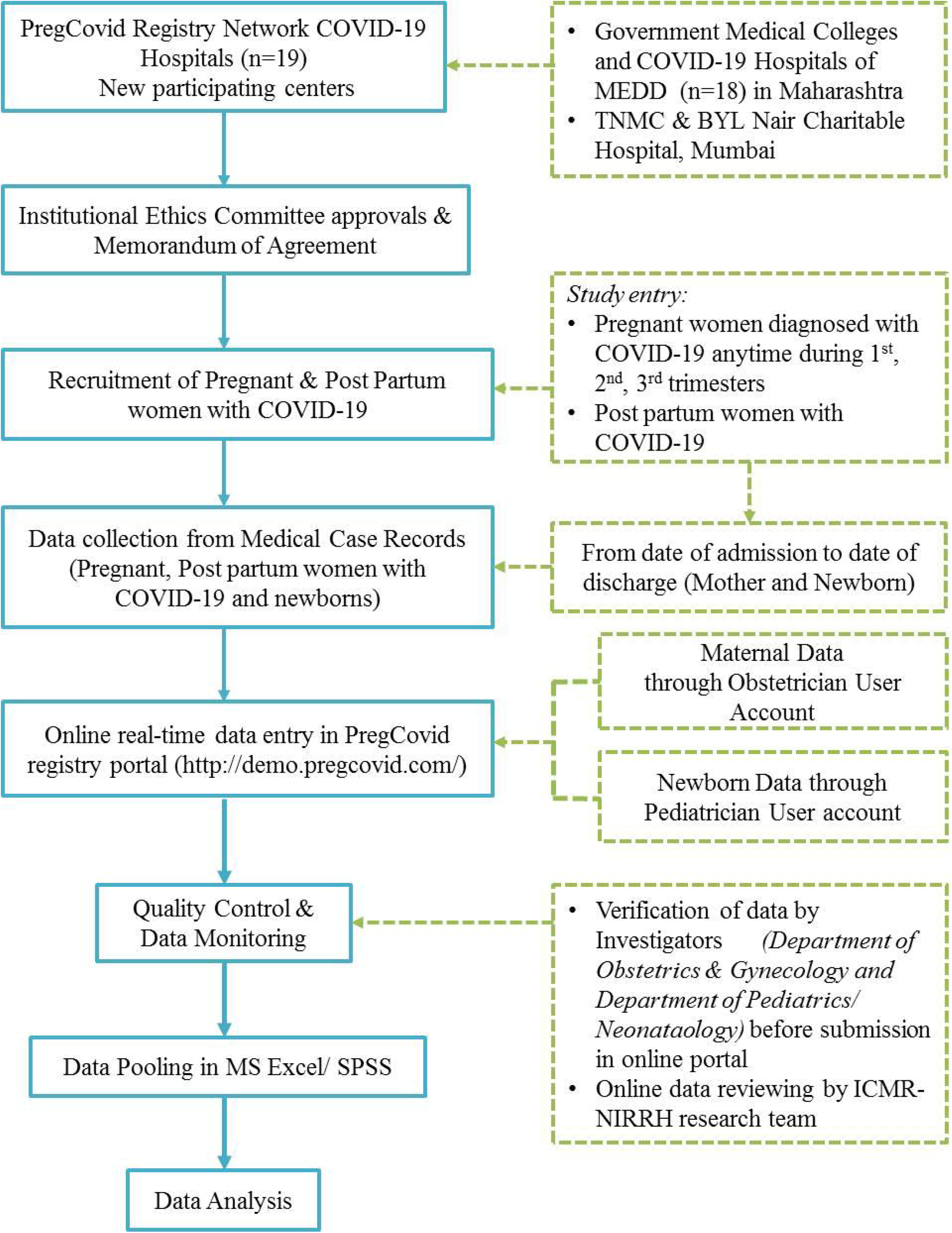
Flowchart showing details on data collection, quality control, data analysis to be used for PregCovid Registry

Maharashtra State is the second most populous state in India with 112,374,333 of which male and female are 58,243,056 and 54,131,277 respectively [17]. The first case of COVID-19 in Maharashtra State was detected in Pune on 9^th^ March 2020 and since then Maharashtra is harboring around 30% of the total cases of COVID-19 burden in India [18]. To respond to this public health emergency, phase 1 of the implementation of the registry will be carried out through the 19 participating centers within Maharashtra.

The registry is open to others who wish to become a part of this network. The approaching centers must contact the Coordinator or the Principal Investigator of the registry at ICMR-NIRRH. They will need to obtain approval from their Institutional Ethics Committee and sign a Memorandum of Agreement with ICMR-NIRRH, Mumbai. For further details, the centers can visit the PregCovid website (https://pregcovid.com/for-covid19-hospitals/).

### Timelines

The registry will collect anonymized real-time data of pregnant women diagnosed with COVID-19 and admitted until May 2022. The date is extendable depending on the duration of the pandemic. Interim analysis may be conducted at the discretion of the investigators in consultation with other stakeholders. The final analysis will be done after collecting all the data.

### Study instruments

The registry is implemented through an app-based Electronic Patient Record (EPR) which is shared with participating centers and the site Principal Investigator (PI) will be responsible for data entry. Software is developed for online real-time data entry. The data will be entered online in the EPR from the original medical records of pregnant and/post-partum women with confirmed COVID-19 infection admitted in COVID-19 hospitals.

### Case record form

The case record form is designed to include socio-demographic, epidemiological, clinical presentations, laboratory investigations, past medical and surgical history, current pregnancy details, pregnancy complications, treatment details, and outcomes. New-born data collection tool includes date and time of delivery, mode of delivery, birth weight, sex of infant, whether livebirth/stillbirth, diagnosis with COVID-19 infection (if any), morbidity, and mortality data (Table 1).

### Training for data entry, monitoring, and quality control

The site PIs and their team will be provided online training for data entry, quality control measures, and accuracy. A training manual and a video of detailed instructions on the use of the software will be provided to all participating study sites. The site PI will ensure that the data is entered in a near real-time manner. The site PIs will monitor the entry of individual data and ensure its accuracy. The team at NIRRH will periodically review the data entered for gaps and discrepancies.

### Data management, Protection of Privacy & Confidentiality

No information will be collected related to the identity of the study participants. The names of the COVID-19 patients will not be entered into any of the documents. None of the COVID-19 patient’s details revealing their identity will be used in any reports and publications arising from this study. The information as per the case record form will be captured electronically in a central database created, maintained, and regularly updated at ICMR-NIRRH. Data entry and data access will be independently controlled and held behind secure firewalls. Data collected from different study sites will be kept separately and only specific individuals within a research team will be able to access and edit the data. The quality of the electronic data will be regularly monitored by the PI and research team at ICMR-NIRRH.

### Statistical analysis

The frequency of each symptom will be calculated. The laboratory data will be analyzed for calculating the frequency of laboratory parameters consistently higher in women with COVID-19. The adverse pregnancy and neonatal outcomes will be analyzed and their frequency will be calculated. Response to treatment will be analyzed for frequency calculation (number of women treated with different treatment regimens). The mother-to-child transmission data will be analyzed from the RT-PCR and/ antibody data of neonatal and maternal samples tested wherever the information is available. Data pooling and statistical analysis will be carried out using MS Excel and SPSS version 26 (SPSS South Asia Pvt. Ltd., Bangalore, India).

### Patient and public involvement

No patients or public were involved in the study design, conduct or reporting of our analysis.

## Discussion

To the best of our knowledge, this is the first registry on pregnant women with COVID-19 in India for capturing data on clinical presentation, obstetrics and neonatal outcomes, response to treatment, and mother to child transmission of SARS-CoV-2 infection. While the initial focus is on Maharashtra, as the pandemic unfolds, we anticipate having membership from different states representative of the Indian population. In its present form, it is geared to generate data of the worst affected state in India in a very systematic manner and its interim analysis would help other states to plan their strategies well in advance to manage pregnant women infected with SARS-CoV-2.

The study will also generate information on the ethnic and socio-demographic determinants of the clinical presentation, and outcomes of pregnant women with COVID-19. Together, this data will aid in planning the strategies to handle the epidemic of COVID-19 and pregnancy. This data is extremely important to develop rational management strategies for protecting pregnant women against possible adverse effects of COVID-19. This information will be also of importance not just for COVID-19 but also provide a broad framework to deal with an entire group of coronaviruses that lead to common respiratory infections in general.

The preliminary findings of the registry data are useful for improved management of pregnancy with COVID-19. ICMR recommends universal testing of pregnant women in India. Accordingly, women residing in clusters/containment areas or large migration gatherings/evacuees centers from hotspot districts in India and presenting in labor or likely to deliver in the next 5 days were recommended to be screened for SARS-CoV-2 [19]. We observed a 12 % prevalence of SARS-CoV-2 in pregnant women with the presence of one symptomatic to every nine asymptomatic pregnant women [13]. These findings of the universal testing strategy of pregnant women are useful for planning strategies to prevent the spread of the virus to newborns, healthcare workers, and others in the community. The study also highlights that pregnant women should be paying special attention to their health. The significance of detecting asymptomatic pregnant women is useful for ensuring safe obstetric and neonatal services and assessing the burden of COVID-19 in the region to plan strategies on strengthening or relaxing mass physical distancing measures.

Detection of dengue and malaria coinfection with COVID-19 in pregnant women is useful for formulating the strategies for pregnant women living in malaria and dengue-endemic zones [20]. Considering the COVID-19 reaching the tribal and rural parts of India, both public and private healthcare systems should be strengthened for diagnosis and appropriate management of co-infections. Documentation of post-partum psychosis in mothers with COVID-19 [21] is useful for creating awareness and providing appropriate mental health care for pregnant and postpartum women with COVID-19.

Globally, there are several National, Regional and International registries being established for pregnant women with COVID-19[22]. The objectives and the protocol of the PregCovid registry are at par with these international registries and will open up an opportunity at the international level for data comparison and assimilation. Eventually, collaborations with an international consortium with global experts having experience in managing COVID-19 pregnant women would be useful for developing a global strategy on COVID-19 in pregnancy.

### Publication plan

Publications will be submitted to international peer-reviewed journals on findings emerged from the data of the PregCovid study. Press releases and articles for lay audiences will also be prepared for important findings.

## Data Availability

Data sharing is not applicable as no datasets were generated and/or analyzed for this study.

## Abbreviations

COVID-19: Coronavirus disease 2019
CTRI: Clinical Trial Registry of India
EPR: Electronic patient record
IRB: Institutional Review Boards
MEDD: Medical Education and Drugs Department
MCGM: Municipal Corporation of Greater Mumbai
MERS-CoV: Middle East Respiratory Syndrome
MoHFW: Ministry of Health and family Welfare
PRIORITY: Pregnancy Coronavirus Outcomes Registry
SARS-CoV: Severe acute respiratory syndrome coronavirus
SARS-CoV-2: Severe acute respiratory syndrome coronavirus 2

## Acknowledgments

The authors sincerely acknowledge the support of the Director-General, Indian Council of Medical Research. Secretary, Medical Education, and Drugs Department; Director, Medical Education, and Drugs Department, Maharashtra; Dean, TNMC, Mumbai & BYL Nair Hospital; are acknowledged. Dr. Vandana Bansal, Dr. Dipti Tandon, Dr. Anushree Patil, Dr. Sudha Rao, Dr. Shakuntala Prabhu, Dr. Pooja Bandekar are sincerely acknowledged. All the Site Investigators and faculty, residents, interns working in the Departments of Obstetrics & Gynecology; Pediatrics and Neonatology of the network of the National Registry of Pregnant women with COVID-19 in India are sincerely acknowledged (PregCovid Registry, CTRI/2020/05/025423).

## Data availability

Data sharing is not applicable as no datasets were generated and/or analyzed for this study.

## Authors’ contributions

Concept and design: RG

Development and validation of data collection instrument: RG, NM, RW, SS,

Pilot testing & validation of software: NM, AB, MP, RW, PH

Development of Training manual, online training: NM, AB, MP, RG, RW

Drafting of the manuscript: RG, NM, DM, SM

Critical revision of the manuscript for important intellectual content: All authors

Administrative and technical support: RG, RW, NM, SM

